# ELISA detection of SARS-CoV-2 antibodies in saliva

**DOI:** 10.1101/2020.08.17.20176594

**Authors:** Melanie A. MacMullan, Albina Ibrayeva, Kylie Trettner, Laura Deming, Sudipta Das, Frances Tran, Jose Ricardo Moreno, Joseph G. Casian, Prithivi Chellamuthu, Jeffrey Kraft, Kenneth Kozak, Fred E. Turner, Vladimir I. Slepnev, Lydia M. Le Page

**Affiliations:** Curative Inc, San Dimas, CA 91773, USA; Mork Family Department of Chemical Engineering and Materials Science, Viterbi School of Engineering, University of Southern California; Eli and Edythe Broad Center for Regenerative Medicine & Stem Cell Research at USC, Department of Stem Cell Biology and Regenerative Medicine, W.M. Keck School of Medicine; Davis School of Gerontology, University of Southern California, Los Angeles, CA 90089, USA; Bridge Institute, Loker Hydrocarbon Research Institute and Department of Chemistry, University of Southern California

## Abstract

To facilitate containment of the COVID-19 pandemic currently active in the United States and across the world, options for easy, non-invasive antibody testing are required. Here we have adapted a commercially available, serum-based ELISA for use with saliva samples, which will enable widespread, affordable testing for patients who experienced this disease.

---

Following the first confirmed case of the 2019 coronavirus (COVID-19) in Wuhan, China on December 2, 2019^1^, cases spread both within China and worldwide, with the World Health Organization declaring COVID-19 a pandemic on March 11, 2020^2^. 89,625 cases have been reported in China as of August 14 2020^3^. Worldwide, as of that same date, more than 20 million cases and over 750,000 confirmed deaths have been reported. Approximately a quarter of these cases and a fifth of these deaths have occurred in the United States, following the first known case in the US reported on 19 January 2020 in Washington State^4^. The continuing, devastating presence of this virus in the US makes it essential that the best tools are developed to understand this disease in as great a population as possible.

Taking into account our current understanding of previous coronaviruses, it is expected that patients with SARS-CoV-2 will develop antibodies against the virus. The emerging literature supports this, although our understanding of the rates of seroconversion and timelines of immunoglobulins M, G, and A is still developing^5^. Antibody detection is typically performed using patient serum, requiring the use of trained phlebotomists and limiting the ability to test a large patient population. Further, a method for self-collection of samples is needed to minimize potential exposure of healthcare personnel during interaction with a patient. As seen with testing for the virus, one sample type has been insufficient and testing per capita has been hugely variable between countries^6^. In the US, the Centers for Disease Control and Protection now recommend options of several upper respiratory tract specimen types^7^ from which to test for the virus. This variety of specimen options provides greater opportunity for widespread testing, and the easiest methods of sampling would enable widespread and frequent testing.

Sensitive antibody tests for other viruses have been successful in dried blood spot^8,9^ and saliva samples^10–12^. Saliva samples are particularly attractive given the ease and non-invasiveness of sampling, allowing for substantial scaling of testing. Initial discussion^13,14^ and testing in small sample sets indicates the potential for using saliva for SARS-CoV2 antibody testing^15^.

In this manuscript we hypothesized that saliva is an acceptable specimen for the detection of SARS-CoV-2 antibodies by standard ELISA. To test this, we first assessed the sensitivity and specificity of two commercially available assays for serum (following manufacturers’ instructions). We then investigated multiple strategies for conversion of these assays for antibody detection in saliva using clinical samples collected from RT-PCR-positive participants who were determined to have antibodies as detected by serum testing.

We conclude that, when following our optimized protocol, it is possible to detect antibodies against SARS-CoV-2 in saliva samples at a sensitivity of 84.2% and a specificity of 100% in the general symptomatic population. If this population is limited to those over the age of 40, sensitivity of 91.5% and specificity of 100% is achievable.

Commercially available SARS-CoV-2 serum-based antibody detection ELISA kits from Gold Standard Diagnostics (GSD) and EuroImmun (EI), which detect the nucleocapsid (N) and spike (S) SARS-CoV-2 structural proteins respectively, were evaluated for their efficacy in detecting IgA and IgG antibodies against the novel coronavirus (SARS-CoV-2) in clinical serum samples **(Fig.1)**. Due to uncertainty about which antibodies are most persistent over time^5^, kits for detecting both IgA and IgG antibodies were evaluated. Kits for detecting IgM antibodies were also considered, but due to a lack of consensus surrounding the persistence of IgM in blood^16^ we chose to pursue IgG and IgA antibodies only. 76 serum samples collected prior to November 2019 (pre-SARS-CoV-2) were used as a negative control panel to evaluate specificity for both IgG and IgA kits from each manufacturer. We determined that both GSD kits and the EI IgG kit were 100% specific, while the EI IgA kit was 92% specific. We then also tested these kits for sensitivity against clinical serum samples collected between April and July 2020.

**Figure 1:**
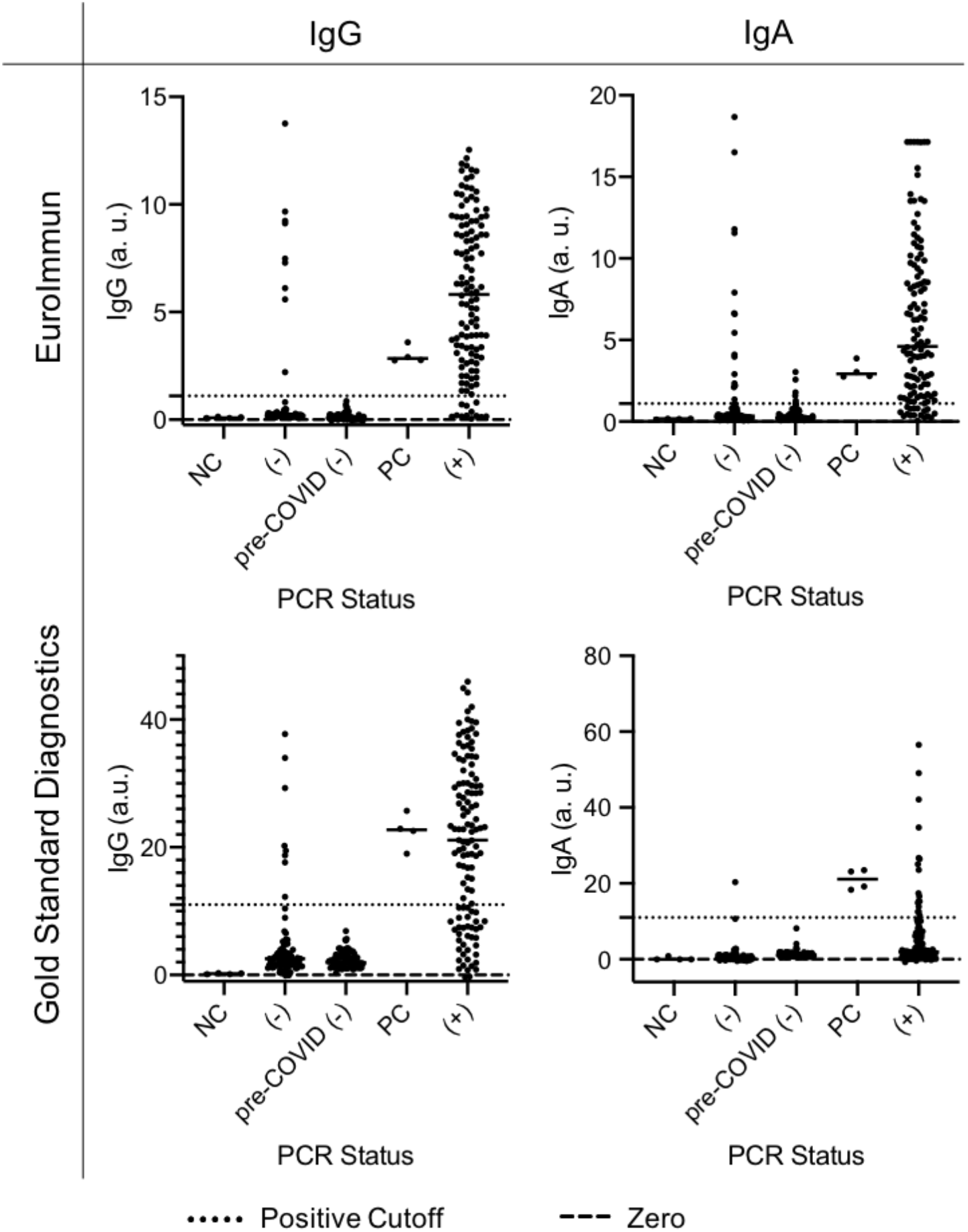
Detection of IgG or IgA antibodies against SARS-CoV-2 in serum samples collected prior to the spread of COVID-19 (pre-Nov. 2019), and people who previously tested negative (-) or positive (+) for COVID-19 using Curative's oral fluid PCR test. The positive cutoff values represented by the dotted lines were provided by the manufacturer. NC = negative control; PC = positive control

Because of limited and confounding existing data on asymptomatic patients and the increased sensitivity 21 days post-symptom onset reported by the ELISA manufacturers, we chose to only investigate samples from symptomatic participants collected more than 21 days post-symptom onset. We tested serum collected from 123 symptomatic, PCR-positive patients and from 83 PCR-negative patients on both IgG and IgA ELISA kits from each manufacturer **(Fig. 1)**. Based on the virus positive patient samples, EI IgG and IgA kits were 90 and 86 percent sensitive respectively and GSD IgG and IgA kits were 69 and 15 percent sensitive respectively based on cutoff values for serum detection supplied by the manufacturers. Nine participants who had tested PCR-negative for RNA derived from SARS-CoV-2 were positive for IgG antibodies against the virus based on both EI and GSD antibody detection kits, suggesting that these patients were likely infected by COVID-19 prior to their RT-PCR test **(Supp. Fig. 1)**.

Our objective was to evaluate the ability of these tests to detect antibodies in saliva in addition to detecting antibodies in serum samples. We selected the EI IgG kit for our saliva sample optimization experiments based on its superior performance in our clinical serum sample testing. All clinical study participants provided oral fluid for viral PCR testing and saliva and serum samples for antibody testing. Saliva samples were collected using devices as described in the Methods section. We selected saliva samples from COVID-19 positive participants (as determined by RT-PCR) that were also positive for antibodies in serum, and samples from COVID-19 negative participants (as determined by RT-PCR) that were also negative for antibodies in serum. Comparing two different devices in a set of saliva samples (**Supp. Fig. 2)** showed that a mouthwash (inhouse formulation, detailed in Methods section) yielded 100% sensitive and specific results for antibody detection. Increasing our sample size to 50 positive and 33 negative samples reduced the specificity to 93.6% and the sensitivity to 84.0% (AUC = 0.946) (**Fig. 2A**). Based on this data, we further optimized the sample preparation and ELISA processes.

**Figure 2:**
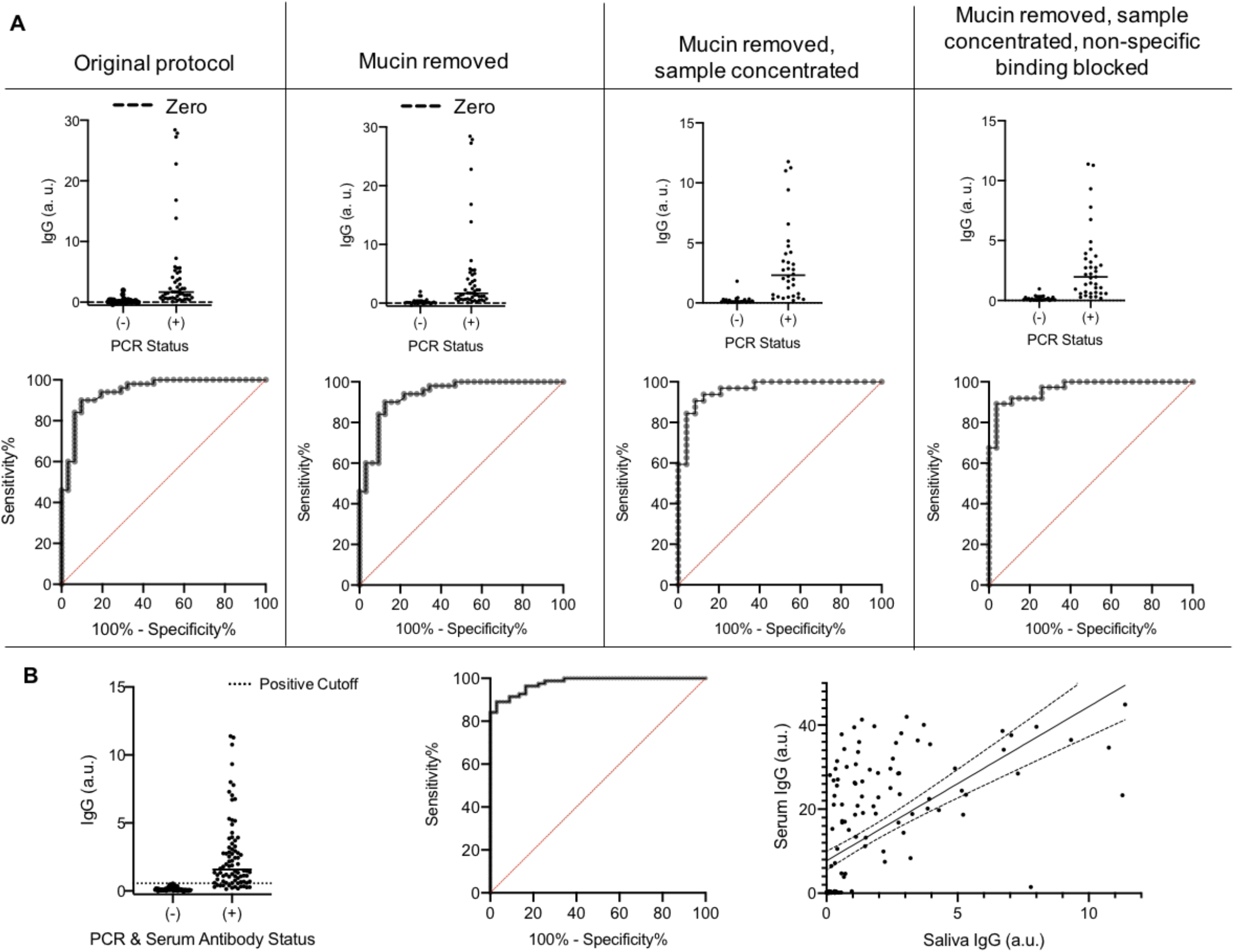
(A) Sample conditions tested for optimized detection of anti-SARS-CoV-2 antibodies in mouthwash saliva samples, in comparison to the original protocol (AUC = 0.946); mucin removed (AUC = 0.935), mucin removed and sample concentrated (AUC = 0.962), and mucin removed, sample concentrated, and non-specific binding blocked (AUC = 0.965) (B) IgG was detected using the optimized protocol in a large sample set where positive and negative status was determined by both PCR and serum antibody status, providing 100% specificity with 84.2% sensitivity (AUC = 0.979). A Pearson correlation showed significant correlation between serum and saliva IgG (R = 0.596, p < 0.0001). Line of best fit is displayed with 95% confidence intervals.

First, centrifuging the samples to remove a large pellet of mucin, a common practice among studies of whole saliva samples^12,17^, gave a sensitivity of 84.0% and specificity of 90.6% (AUC = 0.935). Second, the addition of a concentration step (Amicon Ultra Centrifugal Filters) improved sensitivity to 84.4% and specificity to 95.8% (AUC = 0.962). Finally, blocking the ELISA plate with a buffer containing TBS-Tween and 5% BSA for 30 minutes before adding sample yielded an 89.2% sensitivity and 96.3% specificity (AUC = 0.965) using a positivity cutoff of 0.40; this final protocol yielded the best sensitivity and specificity for this set of mouthwash samples **(Fig. 2A)**.

To determine whether the test remained sensitive and specific in an expanded population, we applied this combined protocol (i.e. centrifugation, concentration, and blocking of the ELISA plate) to a larger mouthwash sample group of 82 positive and 67 negative samples. In this population, our test was 84.2% sensitive and 100% specific (AUC = 0.979) using a positivity threshold of 0.56 **(Fig. 2B)**. Antibody detection in saliva and serum showed a weak but statistically significant correlation (R = 0.596, p< 0.0001). **(Fig. 2B)**.

Given our cohort of clinical samples (demographics in **Fig. 3A**), we wanted to determine if factors linked to COVID-19 susceptibility impacted our sensitivity and specificity. By stratifying the data based on patient demographics of age, sex, and days elapsed since onset of symptoms for sample collection **(Supp. Table 1)**, we found that testing samples only from those over 40 years of age further improved our saliva-based antibody detection test **(Fig. 3B)**. This sample set gave a sensitivity of 91.5% and specificity of 100% (AUC = 0.989, **Fig. 3C**). This optimized test could be used on populations of patients of 40 years or more of age, where we might suggest a borderline range between IgG ratios of 0.40 and 0.55, and consider positive tests having an IgG ratio greater than 0.56.

**Figure 3:**
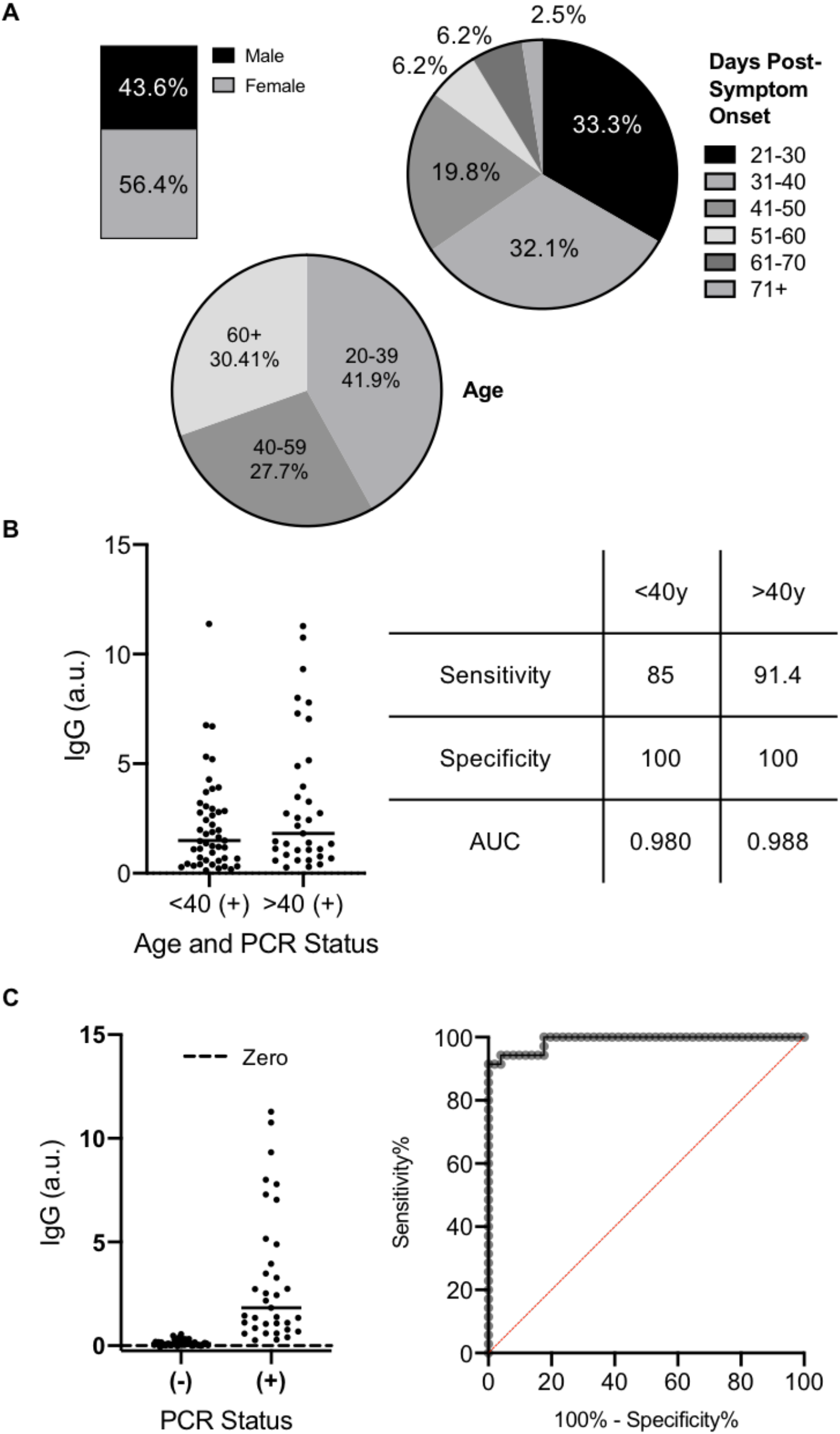
(A) Visualization of the gender (n = 149), age (n = 149), and days post symptom onset (n = 85) of our cohort of clinical samples. (B) Stratifying by age impacted the sensitivity and specificity of our optimized saliva protocol. (C) When limited to sampling people over 40 years of age, 91.5% sensitivity and 100% specificity (AUC = 0.988) can be achieved.

In conclusion, we have shown that testing for IgG against SARS-CoV2 is possible in saliva samples, providing an easy, noninvasive option for detection of prior infection. Further optimization and increasing the cohort size may improve this method to the levels recommended by the FDA guidance (that is, better than 90% sensitivity and 100% specificity). Further work could describe results in asymptomatic COVID-19 positive people, estimated to be as high as 40% of cases^18^.

Development of this specific, sensitive antibody test from non-invasive samples which require no exposure of healthcare workers to potentially infectious environments may be very valuable. There is currently no documented reinfection in the US, which suggests immunity may be protective on the order of months. Antibody testing could provide significant information for countries interested in establishing the reach of the disease in order to help make policy decisions about reopening towns, cities, and states.

## Methods

Human serum and saliva sample collection.

### Pre-COVID-19 serum samples

These samples were purchased from Cureline (Brisbane, CA), and were collected before September 2019 from healthy adults in the USA.

### Post-COVID-19 serum and saliva samples

Clinical samples were collected under UCLA Institutional Review Board approved study protocol IRB#20–000703. The UCLA IRB determined the protocol was minimal risk and verbal informed consent was sufficient for the research under 45 CFR 46.117(c)(2). The study team complied with all UCLA policies and procedures, as well as with all applicable Federal, State, and local laws regarding the protection of human subjects in research as stated in the approved IRB.

For this study, we worked with four specimens obtained: oral fluid for viral PCR testing, blood via venipuncture, and two saliva specimens.

#### Oral fluid swab for viral RNA

This was obtained according to the reported guidelines for Curative’s oral fluid COVID-19 test. Briefly, participants coughed hard three times while shielding their cough via mask and/or coughing into the crook of their elbow. They then swabbed the inside of their cheeks, along the top and bottom gums, under the tongue, and finally on the tongue, to gather a sufficient amount of saliva. Swabs were placed in a tube containing RNA shield and transported at room temperature before laboratory processing as described^19^. Positive for viral RNA was determined as below 35 cycle threshold (CT).

#### Blood sampling

Participants underwent a standard venipuncture procedure. Briefly, licensed phlebotomists collected a maximum of 15ml whole blood into 3 red-top SST tubes (Becton-Dickinson, cat. number 367988). Once collected, the sample was left at ambient temperature for 30–60 minutes to coagulate, then was centrifuged at 2200–2500 rpm for 15 minutes at room temperature. Samples were then placed on ice until delivered to the laboratory site where the serum was aliquoted to appropriate volumes for storage at –80 ºC until use.

#### Orasure saliva sample collection

Orasure oral specimen collection devices (catalog number 3001–2870, Orasure, USA) were used as instructed^20^. The pad was brushed briefly on the lower gums and then held between the gum and the cheek for 2–5 minutes. The pad was then placed into the storage tube, with the provided storage solution. Samples were kept on ice until they reached the lab. The samples were processed as recommended by the manufacturer^21^ before being aliquoted and stored at –80°C until use.

#### Mouthwash saliva samples

Mouthwash was made in-house by adding 3% Sodium Chloride (15g), 0.2% Citric Acid (1g), 0.075% Sodium Benzoate (0.375g), 0.075% Potassium Sorbate (0.375g) to 500mL of ddH_2_O. The solution was then autoclaved (121ºC for 20 minutes) before pH adjustment to pH = 6.5 with 0.1M of NaOH. 4ml aliquots were then used to vigorously rinse the mouth for 1–2 minutes before being collected. Samples were kept on ice until they reached the laboratory, where they were aliquoted (avoiding any large particulates in the liquid) and stored at –80°C until use.

### Serum ELISA

EuroImmun SARS-CoV-2 IgA and FDA EUA IgG ELISAs^22^ for serum (cat. numbers EI 2606–9620 IgA, EI 2606–9620 IgG, EuroImmun, New Jersey, USA) targeting spike (S) protein were run according to the manufacturer provided protocol^23^ on the Thunderbolt (Gold Standard Diagnostics, Davis, CA) automated instrument. Briefly, serum was diluted 1:101 in each well with the provided sample buffer and then incubated at 37ºC for 1 hour. Sample wells were washed three times with a provided wash buffer (10X dilution with ddH_2_O, 0.35ml per well), before the provided conjugate solution was added (0.1ml per well) and incubated at 37ºC for 30 minutes. After a second wash step, the provided substrate solution was added (0.1ml per well) and incubated at ambient temperature for 30 minutes. The provided stop solution was then added (0.1ml per well) and absorbance of sample wells measured immediately at 450 nm and 630 nm, with output reports generated with optical density (O.D.) at 630nm subtracted from O.D. at 450nm. Data were then analyzed as recommended by the manufacturer and results reported as a ratio(**Equation 1**).

**Equation 1**. Determination of sample absorbance ratio based on sample O.D. divided by the averaged O.D. of the calibrators.

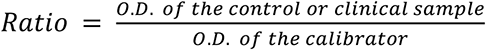

Gold Standard SARS-CoV-2 IgG and IgA ELISAs for serum (cat. numbers GSD01–1029 IgA, GSD01–1028 IgG, Gold Standard Diagnostics, Davis, USA) targeting nucleocapsid (N) protein were run according to the manufacturer provided protocol on the Thunderbolt (Gold Standard Diagnostics, Davis, CA) automated instrument. Briefly, serum was diluted 1:101 in each well with the provided sample buffer and then incubated at room temperature for 30 minutes. Sample wells were washed three times with a provided wash buffer (20X dilution with ddH_2_O, 0.3ml per well), before the provided conjugate solution was added (0.1 ml per well) and incubated at ambient temperature for 30 minutes. After a second wash step, the provided substrate solution was added(0.1ml per well) and incubated at ambient temperature for 30 minutes. The provided stop solution was then added (0.05ml per well) and absorbance of sample wells measured immediately at 450 nm and 630 nm, with output reports generated with optical density (O.D.) 630nm subtracted from O.D. at 450nm. Data were then analyzed as recommended based on a correction factor (specific to each kit) and mathematical formula provided by the manufacturer (**Equation 2, 3**).

**Equation 2**. Determination of sample positivity cutoff value as an average of the calibrator values multiplied by a lot specific correction factor.

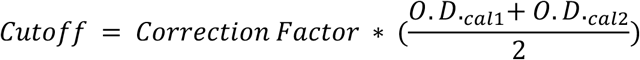

**Equation 3**. Determination of the antibody units as defined by the manufacturer by dividing sample O.D. by the positivity cutoff value and multiplying by 10.

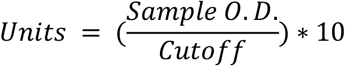

### Saliva ELISA optimizations

#### Centrifugation of samples

1ml of mouthwash samples were centrifuged at 15,000 rpm for 10 minutes (4ºC). The supernatant was then immediately transferred to a clean tube.

#### Concentration of samples

300ul of supernatant was placed in Amicon Ultra Centrifugal Filters (UFC5199BK, Millipore Sigma, USA) and centrifuged at 4ºC, 15,000rpm for 8 minutes. The concentrated sample was then recovered via a second centrifugation step (4ºC, 15,000 rpm for 5 minutes); this procedure produced ∼50ul of concentrated sample. 50ul of a blocking buffer prepared in house (5% BSA in TBS with 0.5% Tween 20) was then added before the total volume (∼100ul) was placed in each well of the ELISA plate, and the protocol run as described above.

#### Blocking of ELISA plate

Prior to concentrated sample loading, 25uL of the in house blocking buffer (5% BSA in TBS with 0.5% Tween 20) was added to each well. Wells were incubated with a blocking buffer for 30 minutes at room temperature before adding sample to begin the ELISA.

### Statistical analysis

ROC curves were generated in GraphPad Prism (GraphPad Prism Version 8.4.3, San Diego, USA), with a 95% confidence interval. Area under the curves was also calculated. Correlation between serum and saliva IgG values were calculated using a Pearson correlation computing R between the two datasets, with a 95% confidence interval.

## Data Availability

The data that support the findings of this study are available from the authors upon reasonable request.

## Acknowledgements

The authors would like to thank Suzana Car, Joseph Kapcia III, Cedie Bagos, Aaron Angel, Marilisa Santa-Cruz and Matthew Geluz.

## Author Contributions

M.M and A.I. designed and ran experiments, analyzed and interpreted data, and drafted the manuscript, K.T. analyzed and interpreted data and edited the manuscript, F.T., J.R.M., J.C. and P.C. edited the manuscript, L.D., S.D, J.K., K.K., F.E.T., V.S. and L.M.L.P. designed experiments, interpreted data, and drafted the manuscript, and F.E.T, V.S. and L.M.L.P conceptualized the project.

